# Impact of the World Inflammatory Bowel Disease Day and Crohn’s & Colitis Awareness Week on Population Interest between 2016 and 2020: Google Trends Analysis

**DOI:** 10.1101/2021.08.01.21261456

**Authors:** Krixie Silangcruz, Yoshito Nishimura, Torrey Czech, Nobuhiko Kimura, Hideharu Hagiya, Toshihiro Koyama, Fumio Otsuka

## Abstract

**Background:** More than 6 million people are affected by inflammatory bowel disease (IBD) globally. World IBD Day (WID; May 19) and Crohn’s and Colitis Awareness Week (CCAW; December 1–7) occur yearly as national health observances to raise public awareness of IBD, but their effects are unclear.

**Methods:** This study evaluates the impact of WID and CCAW on the public awareness of IBD in the United States (US) and worldwide from 2016 to 2020 using the relative search volume (RSV) of “IBD,” “Ulcerative colitis,” and “Crohn’s disease” in Google Trends (GT). To identify significant timepoints of trend changes (joinpoints), we performed Joinpoint regression analysis.

**Results:** No joinpoints were noted around the time of WID or CCAW during the study period in the search results of the US. Worldwide, joinpoints were noted around WID in 2020 with the search for “IBD” and around CAAW in 2017 and 2019 to search for “ulcerative colitis.” However, the extents of trend changes were modest without statistically significant increases.

**Conclusions:** WID and CCAD may not have worked as expected to raise public awareness of IBD. Additional measures are necessary to help raise awareness of IBD related to the health observances.

**Summary:** World IBD Day and Crohn’s and Colitis Awareness Week are key health observances related to IBD. These observances, however, might not have been effective in raising public awareness of IBD in the US and worldwide, according to Google Trends analysis.

## Introduction

Inflammatory bowel disease (IBD) is a global disease with an increasing prevalence in newly industrialized countries and rising cases documented in every continent^1^. Recent systematic reviews have demonstrated that IBD is occurring at increasing rates in such countries^2^. Globally in 2017, there were 6.8 million cases of IBD with an increased age-standardized prevalence rate from 1990 to 2017^2^. Within the United States, it was estimated that more than a million adult Americans had IBD^3, 4^.

Research into Inflammatory bowel disease, however, is largely under-represented despite its prevalence due to a multifactorial nature of the disease^5^. Global efforts have been made to raise awareness for IBD. In 2010, World IBD Day (WID) was created by the European Federation of Crohn’s and Ulcerative Colitis Associations and patient organizations to increase IBD awareness and provide education about IBD to the public^6, 7^. Similarly, Crohn’s and Colitis Awareness Week (CCAW) was created by an US Senate resolution in 2011, with goals of encouraging all people in the US to engage in activities aimed at raising awareness of IBD among the general public^8^.

Disease awareness and health promotion campaigns are created to increase public health education, awareness and, ultimately, change behavior^9^. Approximately 200 health awareness days, weeks, or months are on the US National Health Observances calendar^10^, and nearly 70% have been introduced after 2005. Despite the increasing number of awareness initiatives, there is a lack of data regarding evidence of their effectiveness and impact^11^. This lack of data highlights the need for greater evaluation and quantifiable metrics to determine the impact of health behaviors on a global scale.

Because online searches are a predominant source of access to health-awareness-related information, internet searches are a reflection of engagement between the public and resources which increase disease awareness. Searches are individual proxies for public disease awareness and provide insight into the effect of dissemination of information via global public health days and weeks. Google Trends (GT) is a novel, open-source, freely accessible resource that allows researchers to interact with search tools to study health phenomena^12^. An analysis regarding the efficacy and public health behaviors which resulted from IBD awareness initiaves has not been done previously. We aim to analyze the relationship between the WID or CCAW, and public health awareness for IBD through GT data.

## Methods

### Data source

GT is a data source generated from the total Google search data^13^. The data are available to the public, and GT has been used in multiple social, public health, or global health research to dig into the public attention^14-25^. Surrogate of the public attention in GT is the relative popularity of specific search terms or topics in a certain category (for example, “health”), place, and time range. The relative popularity is defined as a relative search volume (RSV) with a scale of 0–100 (0 being the lowest popularity)^14, 19-21^. The RSV correlates with how popular the terms are at a certain time point.

### Search Input

**Fig. 1** is a summary of our search strategy with GT. We followed protocols noted by previous studies^17, 19, 21^. We chose [Inflammatory bowel disease], [Ulcerative colitis], and [Crohn’s disease] as search inputs. The location of the search included “United States” and “Worldwide.”

**Figure 1.**
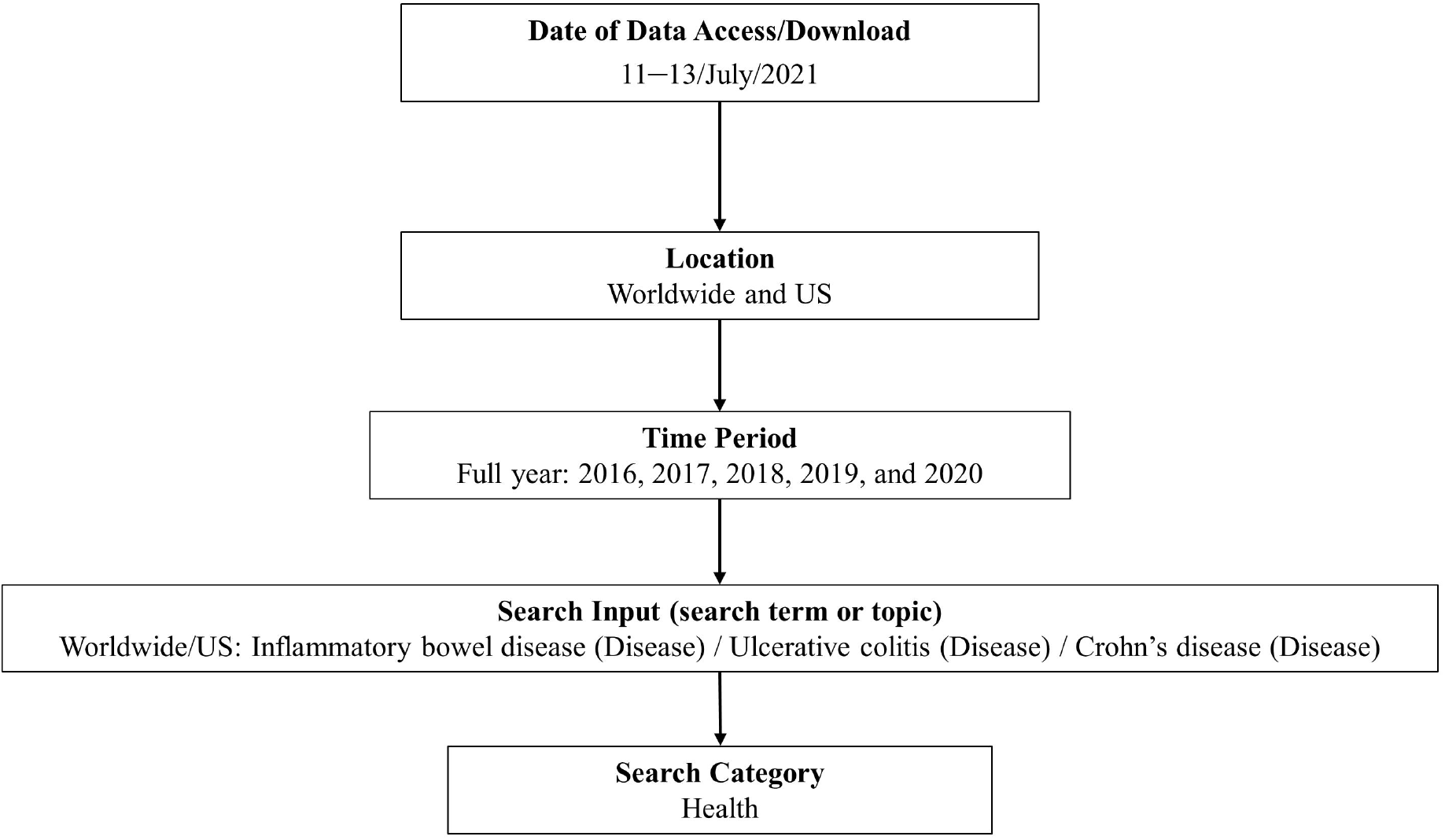
Searching flow using Google Trends.

### Search Variables

To specifically obtain the popularity of the disease-related search inputs, all searches were done with a ‘disease’ option in a Health category (with a “disease” option, search volumes of subtopics or relevant themes are included). We chose each full year from 2016 to 2020 as search scales to visualize weekly trends of the RSVs (each year contains 52 or 53 weeks; the WID occurred in the 20^th^ week of 2016 to 2019, and the 19^th^ week of 2020; CCAD occurred in the 48^th^ to 49^th^ week in 2016 to 2019 and the 49^th^ to 50^th^ week in 2020).

### Statistical analyses

We used a joinpoint regression model with the Joinpoint Regression Program (version 4.9.0.0, March 2021, Statistical Research and Applications Branch, National Cancer Institute, USA)^26^ to analyze the RSV data and their time trend. The software enables us to identify time points called joinpoints, where a temporal trend significantly changes. We defined the analysis criteria to look for up to three joinpoints. The weekly percent changes (WPCs) between trend-change points were determined with 95% confidence intervals (CI). The threshold for statistical significance was defined as a *p*-value < 0.05, suggesting the level at which the slope differed from zero.

### Ethical Considerations

The publicly available data published by GT (Google LLC, Mountain View CA, USA) are utilized in the project. The study was approved by the institutional review board of Okayama University Hospital with a waiver for informed consent since the study intended to retrospectively analyze open data (No. 1910-009). All research methods were performed in accordance with relevant guidelines and regulations.

## Results

### Trends in the search volume of “Inflammatory bowel disease”

**Table 1** and **Figure 2** describe trends and trend changes of the weekly RSVs for “Inflammatory bowel disease” in each full year from 2016 to 2020. With respect to the search results in the US, no joinpoints were observed throughout the period. Regarding the search results worldwide, there was a joinpoint at the 45^th^ week in 2019 before which a significant increase in the WPC of 0.2% (95% CI; 0.1–0.4) was observed. In 2020, a joinpoint was noted at the 17^th^ week (3 weeks before WID), after which there was a significant weekly increase in the RSV by 0.3% (*p* < 0.001). Also, the third joinpoint was observed in the 48^th^ week (a week prior to CCAD). However, no joinpoints were noted from 2015 to 2018 around the time of WID or CCAW.

**Table 1.** Trend changes in relative search volumes of “Inflammatory bowel disease”, 2016–2020.

| Country/Year | Period 1 |  | Period 2 |  | Period 3 |  | Period 4 |  |
| --- | --- | --- | --- | --- | --- | --- | --- | --- |
|  | Weeks | WPC (%) [95% CI] | Weeks | WPC (%) [95% CI] | Weeks | WPC (%) [95% CI] | Weeks | WPC (%) [95% CI] |
| US/2016 | 1–52 | –0.1 [–0.4, 0.2] |  |  |  |  |  |  |
| US/2017 | 1–53 | –0.2 [–0.5, 0.1] |  |  |  |  |  |  |
| US/2018 | 1–52 | –0.2 [–0.5, 0.2] |  |  |  |  |  |  |
| US/2019 | 1–52 | –0.3* [–0.6, 0] |  |  |  |  |  |  |
| US/2020 | 1–52 | 0.1 [–0.2, 0.4] |  |  |  |  |  |  |
| Worldwide/2016 | 1–52 | 0 [–0.2, 0.2] |  |  |  |  |  |  |
| Worldwide/2017 | 1–53 | 0 [–0.2, 0.1] |  |  |  |  |  |  |
| Worldwide/2018 | 1–52 | 0 [–0.2, 0.2] |  |  |  |  |  |  |
| Worldwide/2019 | 1–45 | 0.2* [0.1, 0.4] | 45–52 | –2.6 [–5.4, 0.3] |  |  |  |  |
| Worldwide/2020 | 1–13 | –2.5* [–3.6, –1.4] | 13–17 | 4.6 [–4.9, 15] | 17–48 | 0.3* [0, 0.6] | 48–52 | –5.4 [–10.9, 0.5] |

\* Significantly different from zero (p <0.05)
Abbreviations: WPC, weekly percentage change, CI; confidence interval
Periods were separated as Period 1–4, when the trend changes were statistically detected in the Joinpoint regression analysis during the study period.

**Figure 2.**
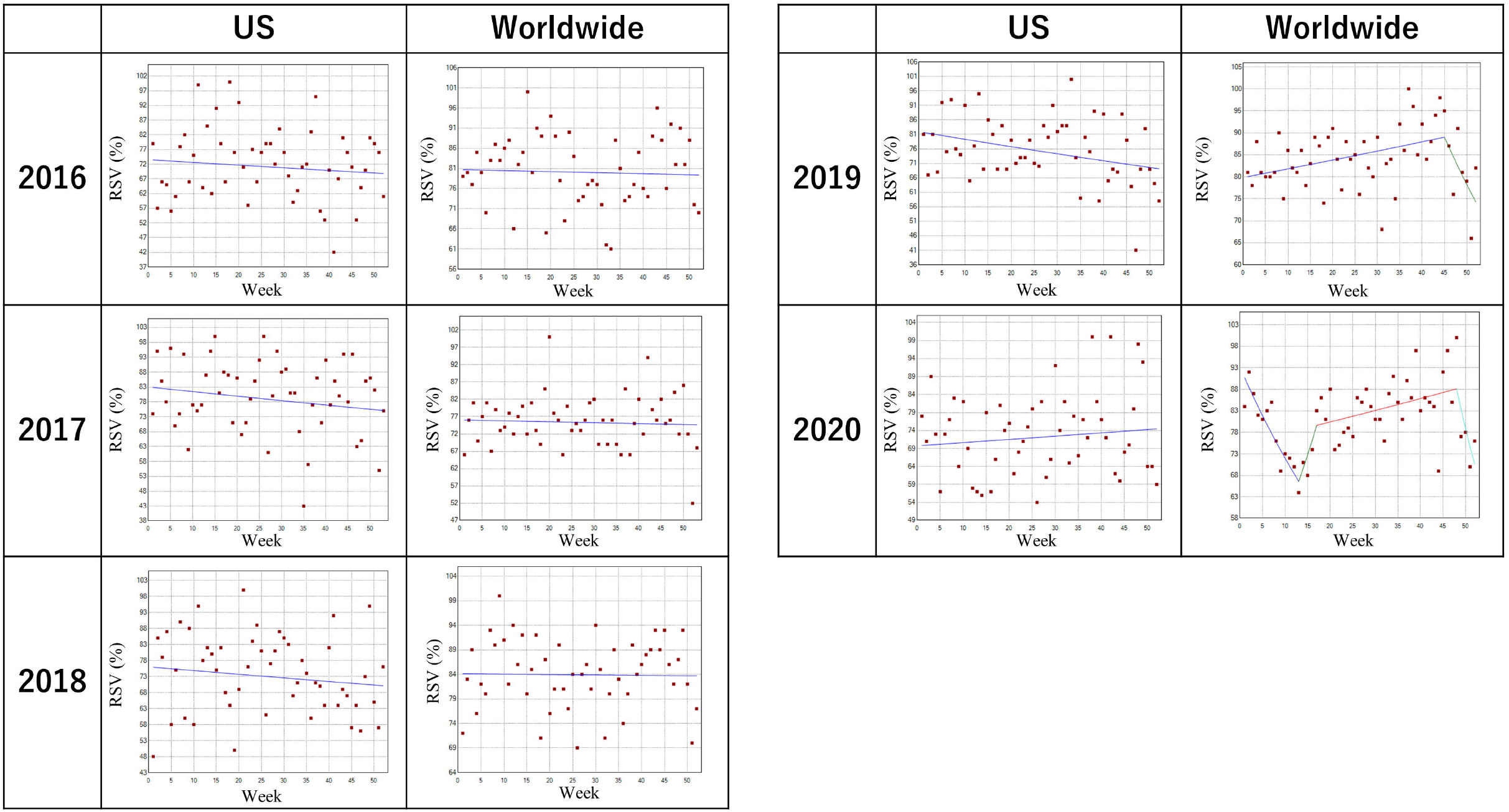
Trends in the relative search volume of “Inflammatory bowel disease,” 2016–2020. Weekly relative search volumes (RSVs) for the search term “Inflammatory bowel disease” are described. World IBD Day (WID) occurred in the 20th week of 2016 to 2019 and the 19th week of 2020; Crohn’s & Colitis Awareness Week (CCAW) occurred in the 48th to 49th week in 2016 to 2019 and the 49th to 50th week in 2020). The number of slopes is determined by the number of joinpoints identified by the analysis. Joinpoints are the time points when statistically significant changes in the linear slopes are noted.

### Trends in the search volume of “Ulcerative colitis”

**Table 2** and **Figure 3** describe trends and trend changes of the weekly RSVs for “Ulcerative colitis” in the designated period. In the search results of the US and worldwide, a big surge was observed at the 3^rd^ week in 2016. In 2020, a joinpoint was noted at the 16^th^ week (4 weeks before WID), after which a non-statistically significant but considerable weekly RSV increase by 3.7% (*p* < 0.001) was observed until 24^th^ week. For worldwide results, there was a prominent joinpoint in the 49^th^ week (CCAW) in 2017. No other joinpoints were observed around the time of WID or CCAW in 2016 or 2018 to 2020.

**Table 2.** Trend changes in relative search volumes of “Ulcerative colitis”, 2016–2020.

| Country/Year | Period 1 |  | Period 2 |  | Period 3 |  |
| --- | --- | --- | --- | --- | --- | --- |
|  | Weeks | WPC (%) [95% CI] | Weeks | WPC (%) [95% CI] | Weeks | WPC (%) [95% CI] |
| US/2016 | 1–3 | 80.2* [40.1, 131.8] | 3–6 | –19.5 [–37.4, 3.6] | 6–52 | –0.2* [–0.4, 0] |
| US/2017 | 1–53 | 0.2 [0, 0.4] |  |  |  |  |
| US/2018 | 1–52 | 0.1 [–0.1, 0.3] |  |  |  |  |
| US/2019 | 1–52 | –0.3* [–0.4, –0.1] |  |  |  |  |
| US/2020 | 1–16 | –2.8* [–4.0, –1.6] | 16–24 | 3.7 [–0.3, 7.8] | 24–52 | –0.5 [–0.9, 0] |
| Worldwide/2016 | 1–52 | –0.3* [–0.6, –0.1] |  |  |  |  |
| Worldwide/2017 | 1–46 | 0.1 [0, 0.2] | 46–49 | 5.7 [–6.5, 19.6] | 49–53 | –7.9* [–11.5, –4.3] |
| Worldwide/2018 | 1–52 | 0 [–0.1, 0.1] |  |  |  |  |
| Worldwide/2019 | 1–46 | 0 [–0.1, 0.1] | 46–52 | –2.1 [–4.5, 0.3] |  |  |
| Worldwide/2020 | 1–52 | 0 [–0.4, 0.3] |  |  |  |  |

\* Significantly different from zero (p <0.05)
Abbreviations: WPC, weekly percentage change, CI; confidence interval
Periods were separated as Period 1–4, when the trend changes were statistically detected in the Joinpoint regression analysis during the study period.

**Figure 3.**
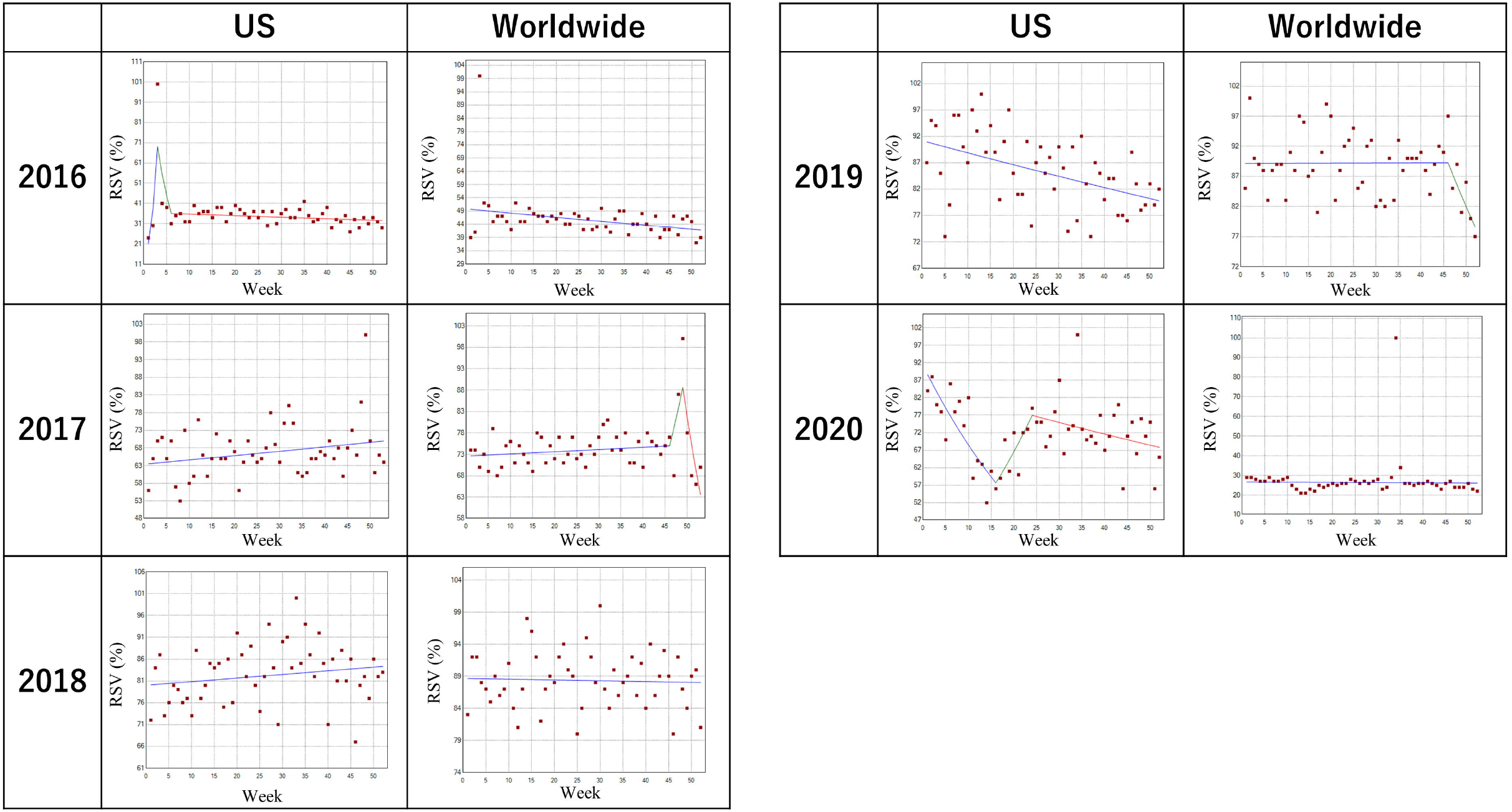
Trends in the relative search volume of “Ulcerative colitis,” 2016–2020. Weekly RSVs for the search term “Ulcerative colitis.” Except for the 16th week (4 weeks before WID) in the US and the 49th week (CCAW) worldwide in 2020, no other joinpoints were noted around the time of WID or CCAW during the designated period.

### Trends in the search volume of “Crohn’s disease”

**Table 3** and **Figure 4** describe trends and trend changes of the weekly RSVs for “Crohn’s disease” in the designated period. Between 2017 and 2019, there was no remarkable trend change in both the US and worldwide. In 2020, joinpoints on the 8^th^ week, the 16^th^ week, and the 24^th^ week in the US. For worldwide, joinpoints were observed on the 10^th^ week, the 14^th^ week, and the 24^th^ week. However, there were no joinpoints around the time of WID or CCAW throughout the period.

**Table 3.**
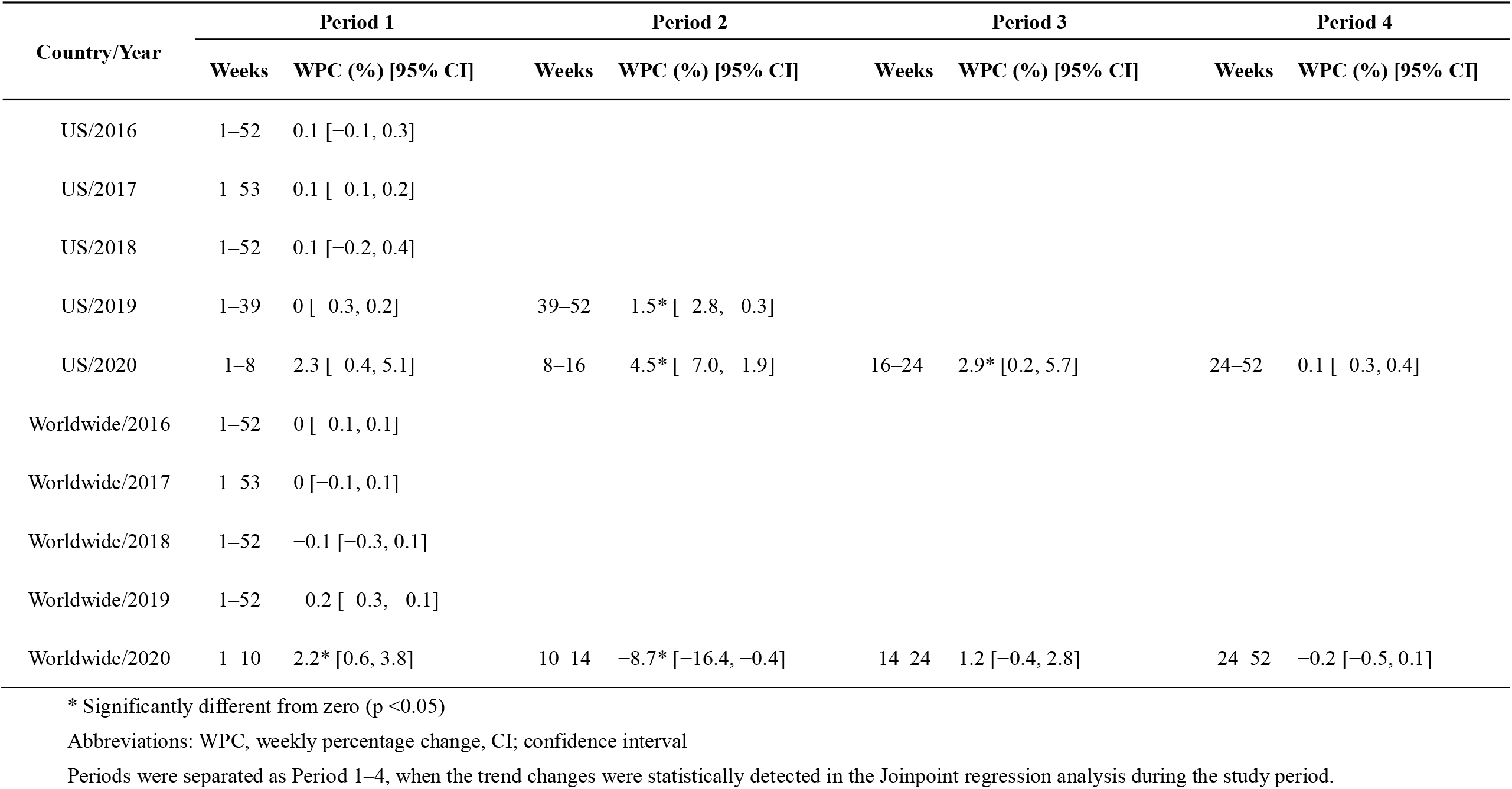
Trend changes in relative search volumes of “Crohn’s disease”, 2016–2020.

**Figure 4.**
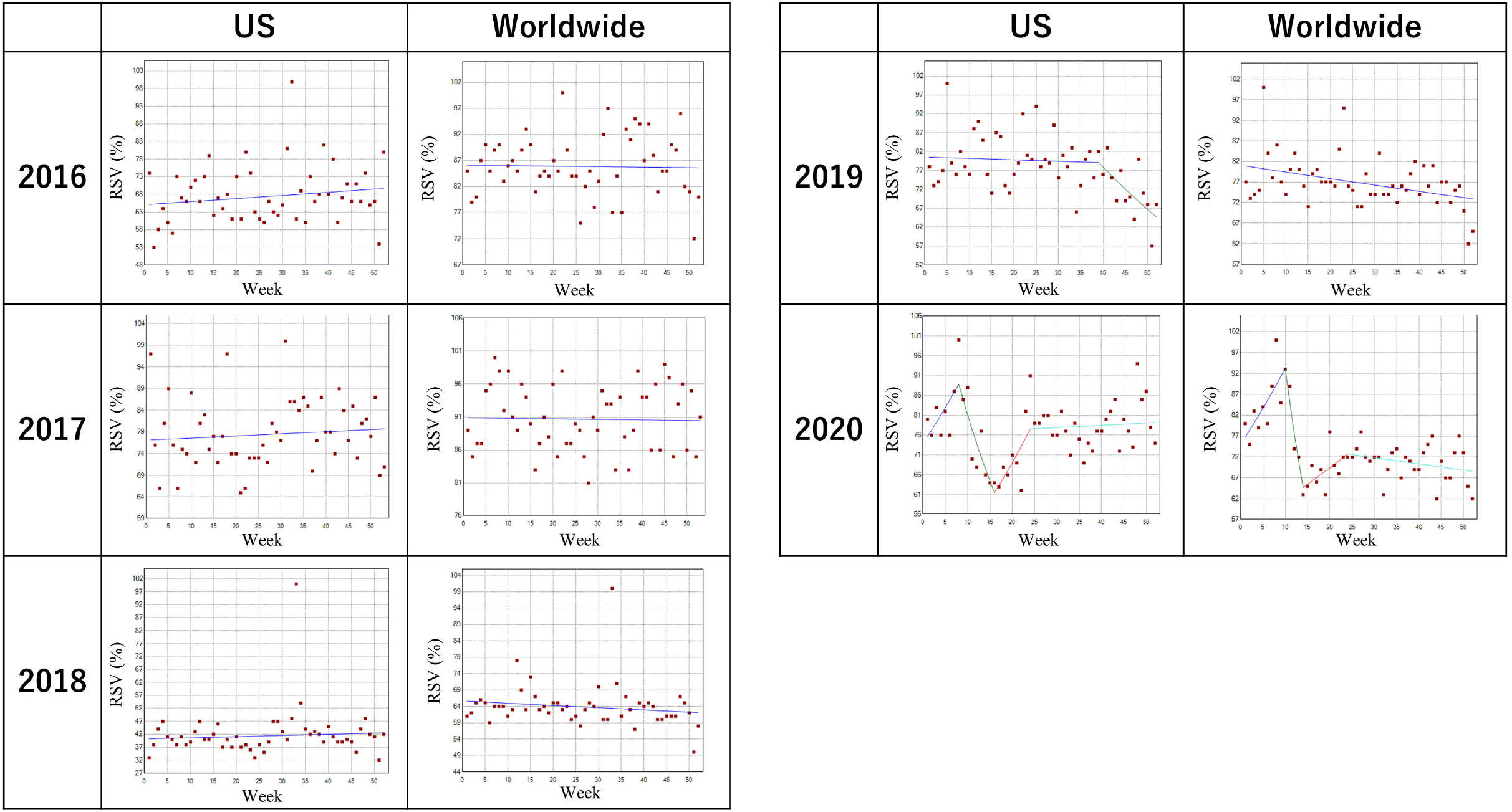
Trends in the relative search volume of “Crohn’s disease,” 2016–2020. Weekly RSVs for the search term “Crohn’s disease.” No joinpoints were noted around the time of WID or CCAW throughout the period.

## Discussion

This study evaluated how the global campaign for promoting IBD, such as WIB and CCAW, affected public awareness using the RSVs of GT data as a surrogate. Although there were several significant joinpoints for IBD, Crohn’s disease, and ulcerative colitis, overall, the present results showed that WID and CCAW might not have significant impact on the public interest in the US and worldwide. Rather, the public interest seems to have been affected by timely topics. For example, at the 3^rd^ week of 2016 when there was a significant increase in the RSV of ulcerative colitis in the US, a famous US singer-songwriter reportedly passed away due to the disease. In March 2020, when significant trend changes were observed in the US and worldwide, a well-known American comedian revealed that he had had Crohn’s disease. Similarly, when the same comedian was featured in a film about a man who has Crohn’s disease in June 2020, there were trend changes both in the US and worldwide (the 24^th^ week). Only in 2017 worldwide, considerable trend changes in the RSVs for ulcerative colitis was noted around CAAW, although the WPC was not statistically significant. Given the rapid increase in the global prevalence of IBD with increasing healthcare costs, raising public awareness of IBD is a pressing global health issue. While one would think that people may be more aware of IBD given the rising number of IBD cases worldwide, more efforts are needed in this aspect.

Since 2020, the dramatic challenges of the coronavirus disease 2019 (COVID-19) pandemic greatly affected our lives, which may have affected the public interests for IBD as well. Because immunocompromised patients may be vulnerable to COVID-19, there were concerns whether IBD patients might be more susceptible to COVID-19 and have worse outcomes^27^. In a cross-sectional questionnaire, patients with IBD were apprehensive about the COVID-19 pandemic, as they felt more vulnerable to COVID-19 due to their condition and their immunosuppressive therapies including biologics. Many patients also felt disturbed, tense when thinking of the infection, and depressed^28^. To provide solid supports for IBD patients during the pandemic, further efforts to increase the public awareness toward the entity is crucial.

The IBD landscape is constantly evolving with the hope as more therapies are discovered, it makes global awareness campaigns even critical. It is crucial to address the essential priorities regarding IBD awareness. There are various ways that campaign organizers can utilize to build awareness and deepen engagement amongst the target audience, not only the general public but also people with IBD, their families, friends, employers, and healthcare providers. Any topic related to treatment always provides positive discussion points and direction to campaign programs. Activities during WID and CCAW and information about the organization itself should be the go-to resource for anything related to IBD. One example of a successful public health awareness campaigns include the annual breast cancer awareness campaign^29^. In order to make public health campaigns more successful, appropriate identification of targets, early involvement of key stakeholders such as celebrities with IBD, and enabling participants to feel part of the campaign using smartphone applications or eHealth platforms may be effective^30^. We encourage societies or organizations associated with WID or CCAW to take actions with features above in mind to improve the effectiveness of these campaigns.

The study’s strength includes that this is the first study to show the public awareness of IBD in the US and worldwide using the GT database. Using the open data, we could quantify the current trends of general awareness in IBD. However, several limitations need to be addressed. First, due to the nature of GT, the present results only included results from those who had internet access and sought health-related information via Google search. Given high internet penetration rates, approximately 90.4% in Northern America and 60.1% worldwide^31^, and high US google search market share, approximately 83%^32^, GT is considered a good surrogate of public awareness. Second, GT lacks full transparency and reproducibility because non-public mathematical assumptions define RSVs. We documented our search strategy in detail to address the limitation. Despite these limitations, our approach is interestingly novel to demonstrate campaign effectiveness or ineffectiveness in public awareness of IBD.

In conclusion, our study suggested that WIB and CCAW might have not been successful to improve public awareness toward IBD using the GT data as a surrogate. There is a need to look deeper into the issues surrounding IBD and how to improve public awareness using these health observances based on good examples including the annual breast cancer awareness campaign.

## Data Availability

The datasets generated and analyzed during the current study are available from the corresponding author on reasonable request.

## Acknowledgments

None.

## Author Contributions

KS, TC, and NK wrote the manuscript. YN proposed the study concept, designed the study, wrote the manuscript, and analyzed the data. HH and TK revised the manuscript critically. FO supervised the research.

